# SARS-CoV-2 Genomic Surveillance Reveals Little Spread Between a Large University Campus and the Surrounding Community

**DOI:** 10.1101/2021.07.19.21260726

**Authors:** Andrew L. Valesano, William J. Fitzsimmons, Christopher N. Blair, Robert J. Woods, Julie Gilbert, Dawn Rudnik, Lindsey Mortenson, Thomas C. Friedrich, David H. O’Connor, Joshua G. Petrie, Emily T. Martin, Adam S. Lauring

**Author notes:** Corresponding author, Adam S. Lauring, MS2 4742C, SPC 1621, 1137 Catherine Street, Ann Arbor, MI 48109.

## Abstract

COVID-19 has had high incidence at institutions of higher education (IHE) in the United States, but the transmission dynamics in these settings are not well understood. It remains unclear to what extent IHE-associated outbreaks have contributed to transmission in nearby communities. We implemented high-density prospective genomic surveillance to investigate these dynamics at the University of Michigan-Ann Arbor and the surrounding community during the Fall 2020 semester (August 16^th^ –November 24^th^). We sequenced complete SARS-CoV-2 genomes from 1659 individuals, including 468 students, representing 20% of cases in students and 25% of total confirmed cases in Washtenaw County over the study interval. Phylogenetic analysis identified over 200 introductions into the student population, most of which were not related to other student cases. There were two prolonged transmission clusters among students that spanned across multiple on-campus residences. However, there were very few genetic descendants of student clusters among non-students during a subsequent November wave of infections in the community. We conclude that outbreaks at the University of Michigan did not significantly contribute to the rise in Washtenaw County COVID-19 incidence during November 2020. These results provide valuable insights into the distinct transmission dynamics of SARS-CoV-2 among IHE populations and surrounding communities.

## Introduction

Institutions of higher education (IHE), such as colleges and universities, have been associated with high incidence of COVID-19 in the United States (Andersen et al., 2021; Leidner, 2021; Vang, 2021). Congregate settings, such as on-campus housing and off-campus social gatherings, have led to large outbreaks of COVID-19 despite mitigation efforts (Currie et al., 2021; Fox, 2021; Weil et al., 2021; Wilson, 2020). It is essential to gain a better understanding of SARS-CoV-2 transmission dynamics surrounding IHE to inform infection prevention strategies (Bradley et al., 2020; Paltiel et al., 2020; Yamey and Walensky, 2020), especially as more contagious variants continue to circulate. In particular, an important but unanswered question is whether IHE-related outbreaks have contributed to transmission in the communities where IHE are geographically located. Some studies have analyzed case counts in areas with IHE, but these approaches are unable to establish transmission linkage between students at IHE and the surrounding community (Bharti et al., 2021; Lu et al., 2021). Counties with large IHE that opened for in-person learning had higher incidence of COVID-19 compared to matched counties that offered remote-only learning (Leidner, 2021). However, it is possible that this reflects transmission only among students without spread into surrounding communities. More direct methods, such as contact tracing and genomic epidemiology, are needed to evaluate whether IHE-related outbreaks have spread into nearby communities.

Virus genome sequencing has been an important epidemiologic tool during the COVID-19 pandemic, enabling the characterization of transmission lineages and their connections across different populations (Martin et al., 2021; Moreno et al., 2020; da Silva Filipe et al., 2021). However, there has not yet been a large-scale genomic surveillance investigation across an entire university in the United States and the surrounding area. Phylogenetic analysis from well-sampled populations can reveal the number of unique introductions of SARS-CoV-2, the growth and persistence of transmission lineages, and the frequency of transmission crossover between groups. An important advantage of genomic surveillance is that it can rule out transmission relatedness between groups that appear to be associated through analyses of case counts and onset dates. The power of genomic surveillance to answer these questions relies on dense, comprehensive sampling in the populations under investigation. Until recently, genomic surveillance sampling density in the United States has been highly uneven across locations (Alpert et al., 2021; Maxmen, 2021). Evaluation of SARS-CoV-2 spread from IHE-related outbreaks into nearby communities requires high-density longitudinal sampling in both populations.

Some recent studies suggest that spillover from outbreaks at IHE occurs infrequently, but the data is not yet conclusive. A recent genome sequencing study described transmission of SARS-CoV-2 from an outbreak among students in La Crosse County, Wisconsin into skilled nursing facilities, resulting in two deaths (Richmond et al., 2020). Other studies have suggested that there has been little spread from outbreaks among IHE students. A seroprevalence study in Pennsylvania indicated that community incidence of COVID-19 remained low throughout a period of high incidence in students (Arnold et al., 2021). Few or no viral descendants of student clusters in Dane County, Wisconsin and Washington State have been detected in genomic surveillance of their respective communities (Currie et al., 2021; Weil et al., 2021). However, because these conclusions rely on the depth and breadth of community surveillance, the extent of community spillover is not completely established. Outbreak investigations typically have higher sampling density compared to community genomic surveillance, which most often ranges from 0.5 – 3% of confirmed cases.

To address this knowledge gap, we conducted prospective, high-density genomic SARS-CoV-2 surveillance during the Fall 2020 semester at the University of Michigan-Ann Arbor and the surrounding area. We sequenced complete SARS-CoV-2 genomes from 1659 individuals, including 468 students. We captured 20% of confirmed cases in University of Michigan-Ann Arbor students and roughly one quarter of confirmed cases in Washtenaw County, where the University of Michigan-Ann Arbor is located. We detected over 200 independent transmission introductions into the student population, including two large transmission lineages that persisted for several weeks across multiple on-campus residences. However, there was very little crossover from student lineages into the community. The largest student-associated lineages waned by mid-November 2020, when community incidence drastically increased. We conclude that outbreaks in students at the University of Michigan-Ann Arbor did not significantly drive the increase in community COVID-19 incidence in southeastern Michigan in November 2020.

## Results

We initiated prospective genomic surveillance in southeastern Michigan in August 2020 with the goal of capturing SARS-CoV-2 transmission dynamics at the University of Michigan-Ann Arbor and the surrounding community in Washtenaw County. We obtained all available residual SARS-CoV-2 positive samples from the University of Michigan Clinical Microbiology Laboratory and the University Health Service (UHS) on a daily basis, starting from August 16^th^, 2020. The University of Michigan Clinical Microbiology Laboratory performs COVID-19 testing for all inpatient and ambulatory clinical settings associated with Michigan Medicine, a large academic medical center that serves over 2.3 million patient clinic visits annually. Testing of students presenting to UHS was either performed on site or sent to the University of Michigan Clinical Microbiology Laboratory. These two specimen sources allowed us to broadly sample the university student population as well as individuals from the community. Unique case identifiers were used to ensure that duplicate specimens were excluded.

### University Setting and Epidemic Course

The COVID-19 epidemic in southeastern Michigan evolved over the course of the Fall 2020 semester. Washtenaw County has a population of over 367,000 and is part of Michigan Public Health Preparedness Region 2 South, which includes Monroe and Wayne counties and the city of Detroit. From August 16^th^ through November 24^th^, there were 6707 laboratory-confirmed COVID-19 cases in Washtenaw County. COVID-19 cases increased in Washtenaw County over the fall semester (Figure 1A), rising from 130 cases during the week of August 23^rd^ (63 daily cases per million) to 1125 cases during the week of November 15^th^ (405 daily cases per million).

**Figure 1:**
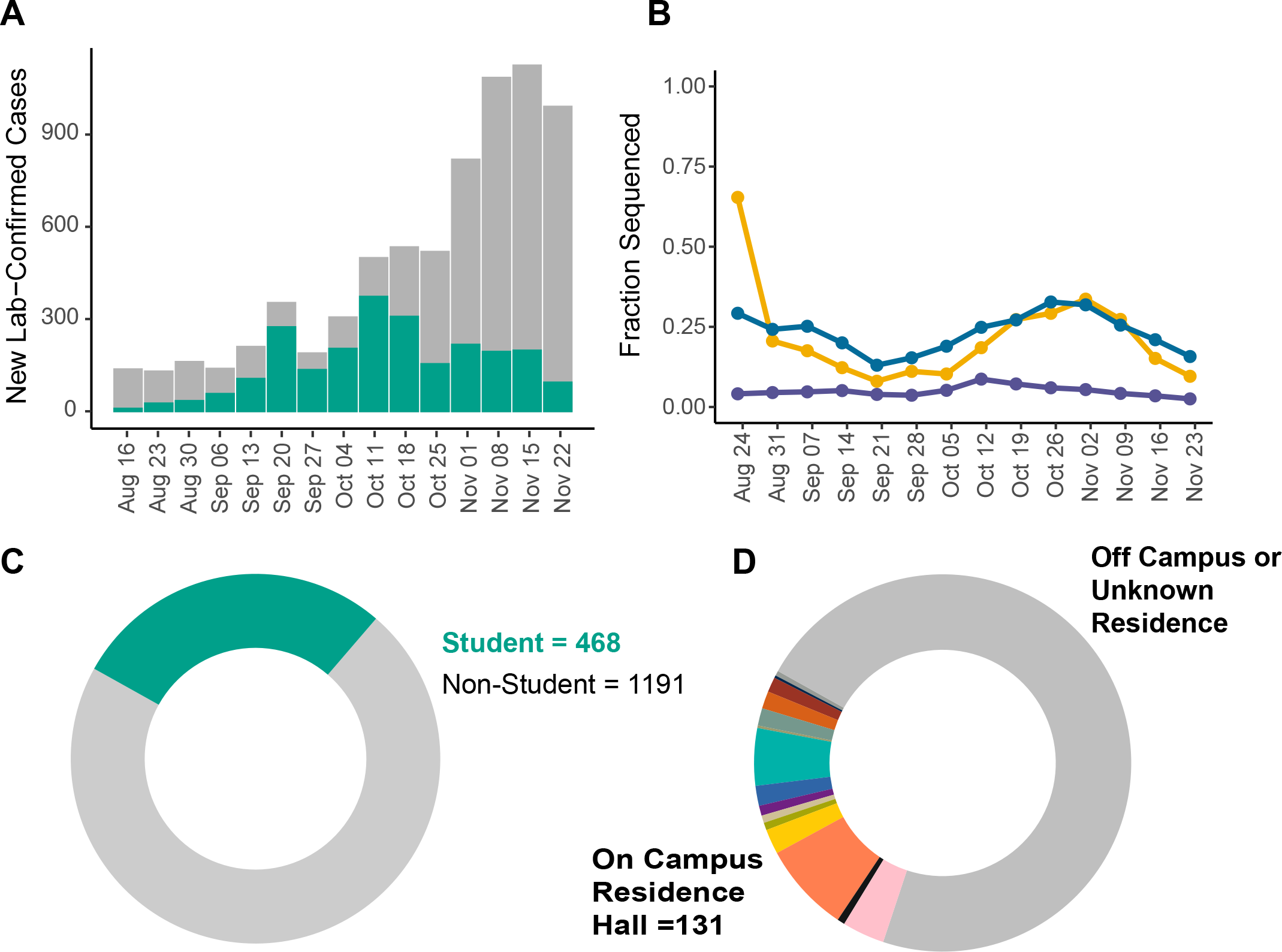
Case curves and sequencing density. (A) New lab-confirmed cases of COVID-19 in Washtenaw County, Michigan from the week of 2020-08-16 through 2020-11-23, displayed by day of symptom onset (as reported by MDHHS). New cases per week are shown on the y-axis and time in weeks on the x-axis. The fraction of new lab-confirmed cases in University of Michigan students is shown in green. (B) Sampling density is displayed as the fraction of new lab-confirmed COVID-19 cases with complete genome sequences (y-axis) per week during the fall term (x-axis). The fraction of student cases sequenced is shown in yellow, all Washtenaw County cases in blue, and all cases in Region 2S in violet (includes Washtenaw, Wayne, and Monroe counties). (C) Genomes sequenced in this study. The number of genomes from cases in students is shown in green and non-students in grey. (D) Of the genomes from cases in students, information on residence hall is displayed (colors).

The University of Michigan-Ann Arbor is a public university with an enrollment of over 47,000 undergraduate, graduate, and professional students. In Fall 2020, the University of Michigan-Ann Arbor opened for on-campus student residence and hybrid in-person and remote learning. In-person instruction was held from August 31^st^ through November 24^th^. Out of 72,798 tests performed from the week of August 16^th^ through the week of November 22^nd^, there were 2374 COVID-19 cases in students, 1064 (44.8%) of which had testing performed at Michigan Medicine laboratories. There was no syndromic case definition for obtaining COVID-19 testing at Michigan Medicine at that time, and testing was available for both symptomatic and asymptomatic cases depending on clinical circumstances. Cases in University of Michigan-Ann Arbor students constituted the majority of cases in Washtenaw County by the end of September 2020 (Figure 1A). In response to this spike, several mitigation efforts were implemented, including stay-in-place orders targeted to specific residence halls, mass testing events, and a broad stay-in-place order for all undergraduates from October 20^th^ –November 3^rd^. The fraction of campus-associated cases in the county declined after mid-October 2020 during a wave of new infections in Washtenaw County in November 2020. The shift in case incidence prompted us to investigate whether the November wave of infections was generated by spillover from outbreaks among students.

### Genomic Surveillance in Southeastern Michigan

We processed all SARS-CoV-2 positive samples tested at UHS or the University of Michigan Clinical Microbiology Laboratory collected between August 16^th^ –November 24^th^ for genome sequencing. November 24^th^ corresponded to the end of in-person instruction for the semester and the peak of the November surge of new cases in Washtenaw County. We assembled complete SARS-CoV-2 genomes from 1659 individuals, representing a median of 24% of the confirmed cases per week from Washtenaw County. For comparison, these genomes represent a median of 4.5% of cases per week from the entire Michigan Region 2 South (Figure 1B). We did not have access to SARS-CoV-2 specimens not tested at Michigan Medicine sites, which limited our sampling density (Figure S1A). Despite this, we sequenced 468 complete genomes from University of Michigan-Ann Arbor students, representing 20% of cases from August 16^th^ through November 24^th^ (Figure 1C, S1B). Of these 468 students, we were able to determine on-campus residences for 131; the on or off campus residence of the remainder are unknown (Figure 1D). The genomes presented here consisted of several different viral clades, mostly Nextstrain clades 20A, 20C, and 20G (Figure S1C). We did not detect any variants of concern that were circulating in other countries at the time, including B.1.1.7, P.1, or B.1.351.

### Transmission Introductions in Students at the University of Michigan-Ann Arbor

We used phylogenetic analysis to characterize the influx of viruses into the student population. We generated a time-calibrated maximum likelihood phylogenetic tree with our sequenced genomes (n = 1659) and additional contextual genomes (See Methods; Figure 2A, S2A). This sequence alignment was a subsampling of data available on GISAID, weighted towards genomes with genetic similarity to those in Michigan. To optimize the inference of transmission lineages in southeastern Michigan, we included all available genomes from Michigan on GISAID that were collected from July – December 2020. We inferred introductions into the student population by discrete trait ancestral reconstruction (see Methods). We inferred traits of ancestral nodes on this time-calibrated phylogenetic tree using a binary model of student vs. non-student, as has been performed in related genomic surveillance studies (Alpert et al., 2021; Lemieux et al., 2020; Müller et al., 2021; Plessis et al., 2021). Genomes from students that shared the same ancestral “student” node were considered part of the same introduced transmission lineage. Genomes from students that were not preceded by a “student” node were considered singleton introductions. We performed the same analysis using a total of ten random subsamples of non-Michigan genomes to verify that our results were not substantially biased by contextual genomes (Lemieux et al., 2020). Due to the large variability in genomic surveillance sampling density from other states in the USA and other countries, we did not attempt to infer the exact geographic sources of transmission introductions into the student population.

**Figure 2:**
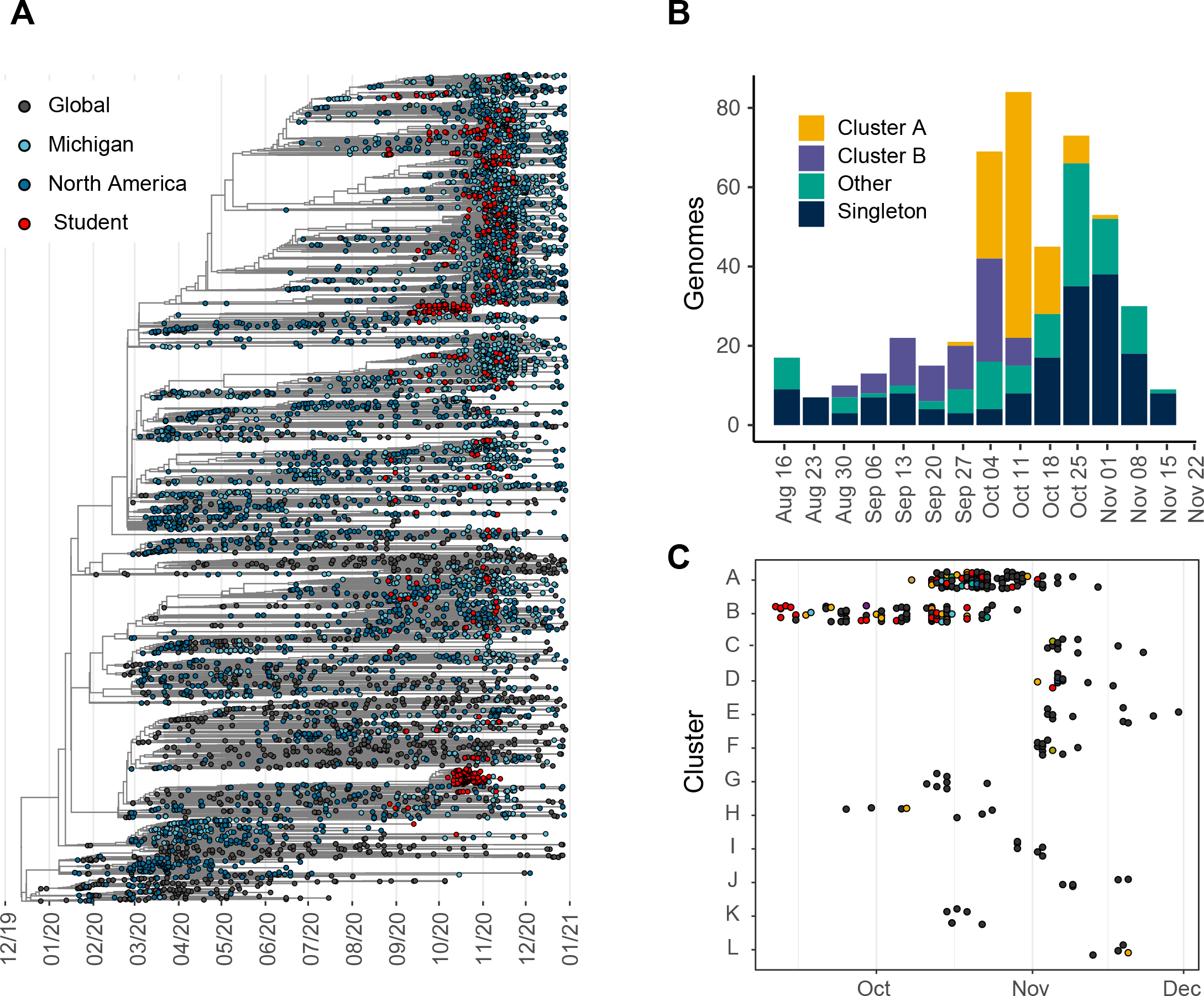
Introductions into the student population. (A) Time-calibrated maximum likelihood phylogenetic tree of 7149 SARS-CoV-2 genomes, including 1657 genomes sequenced for this study. Month is shown on the x-axis. Genomes from students are shown in red, other genomes from Michigan in light blue, other genomes from North America in dark blue, and global genomes in grey. (B) Counts of genomes from students (y-axis) per week during the fall term (x-axis) by the inferred transmission lineage group. Singleton introductions are shown in dark blue, genomes from Cluster A in yellow, genomes from Cluster B in violet, and genomes from smaller clusters (2 – 8 individuals) in teal. (C) Inferred transmission lineages in students, with genomes from each lineage or cluster (y-axis) shown as points with time on the x-axis. Only inferred transmission lineages with ≥5 students are shown. Point colors reflect on-campus residences, if known, as in Figure 1 (unknown residence is black).

Using this approach, we inferred 203 distinct transmission introductions into students (Figure S2B). There were two large transmission lineages in students, which we denote as Cluster A (n = 115 students) and Cluster B (n = 73 students). These lineages were the predominant source of cases in students during the middle of the semester, representing >50% of genomes from students from the week of September 20^th^ through the week of October 18^th^. The rest of the introductions were singletons (n = 171) or small clusters of 2 – 8 students (n = 30 introductions). Small transmission lineages (2 – 8 students) were often short in duration, lasting a median of 3.5 days (IQR 1 – 8 days). We do not have information on social contacts of students within these transmission lineages. We detected singleton cases over the entire course of the Fall 2020 semester. The frequency of singletons increased during the latter half of the semester (Figure 2B), indicating new introductions into the student population rather than spread from older transmission clusters, such as Cluster A and Cluster B. These data suggest a shifting epidemiology in the student population across the semester, characterized by two dominant lineages during the early and middle portions of the semester followed by many small introductions later in the semester as community incidence increased.

We further investigated the two largest inferred lineages, which indicate sustained local transmission of SARS-CoV-2 within the student population. Genomes from Cluster A were part of Nextstrain clade 20B and PANGO lineage B.1.1.304 (TMRCA August 26^th^, 95% CI August 3^rd^ –September 26^th^). Genomes from Cluster B were part of Nextstrain clade 20C and PANGO lineage B.1.593 (TMRCA July 29^th^, 95% CI July 3^rd^ –August 28^th^). We detected cases from Cluster A from October 8^th^ through November 14^th^ and Cluster B from September 11^th^ through October 23^rd^ (Figure 2C). These clusters were genetically distinct from other lineages in our data and identified in all ten independent subsamples (Figure S2C). Cluster A was defined by mutations G4207T, C11471T, G24816C, G25906T, C28399A, and Cluster B was defined by G3013A, T6769C, G12243A, T15567C, C28435T. PANGO lineages B.1.1.304 and B.1.593 are rare in the GISAID database (0.07% and 0.02%, respectively, out of 532,715 genomes from the USA as of June 7^th^, 2021). Among all available genomes from Michigan, these lineages are almost completely unique to this study. Outside of the genomes presented here, B.1.1.304 has been detected in Michigan three times (238 in the USA), and B.1.593 has not been detected elsewhere in Michigan (only twice in the USA). The rarity of these PANGO lineages suggests that these outbreaks among students did not spark significant transmission in southeastern Michigan or in other parts of the country.

We examined the on-campus residence locations for students in these transmission lineages. There was no obvious association with a single residence hall in any transmission lineage in students (Figure 2C). Of the 131 individuals with known on-campus residences, 31% were singletons (n = 40) and 56% (n = 73) were part of Cluster A or B. Individuals from many different residence halls across the University of Michigan-Ann Arbor campus were present in Cluster A and B, including students from nine and seven campus residences, respectively. Of the 188 students in Cluster A and B, 61% (n = 115) did not have an identifiable on-campus residence and likely lived off-campus (Figure 2C, black points). Besides Cluster A and B, there was only one other student lineage that had more than one individual from the same campus residence (2 of 3 students in the lineage were from the same residence). The first individuals detected in Cluster B resided in the same residence hall (Figure 2C), potentially reflecting transmission in a congregate setting. However, without detailed contact tracing to complement the genomic data, it is unclear whether clusters A and B originated in specific residence halls and then spread further among students, or whether the clusters originated in off-campus gatherings. Overall, these data demonstrate local transmission among the student population with intermixing among students from multiple on-campus residences and students residing off-campus.

### Limited Spillover from Student Clusters into the Broader Community

Clusters A and B, defined by PANGO lineages B.1.1.304 and B.1.593, waned by mid-November when new COVID-19 cases increased in Washtenaw County (Figure 3), which suggested that these large, student-dominated lineages did not significantly contribute to the rise in community cases. To further examine the extent of COVID-19 spread from students into the broader southeastern Michigan community, we used the same ancestral trait reconstruction method as above, applying a binary student/non-student model. We enumerated genomes from non-students that descended from an ancestral node that was inferred as “student.” There were very few non-students in the community with genomes that descended from student-associated lineages (n = 53), and most were nested within either Cluster A (n = 18) or Cluster B (n = 11, Figure 4A). A total of 24 non-students descended from the rest of the inferred transmission lineages (2 – 8 students each). Out of the 1191 genomes we sequenced from non-students, 96% (n = 1138) were not genetic descendants of detected clusters in students (Figure 4B). There was a positive association between the size of a cluster among students and the number of non-student descendants (Figure 4C). This means that although the total number of non-student descendants was low, larger transmission lineages among students resulted in more spread to the community than smaller lineages. The median age of non-students descending from Cluster A and B was 47 years (IQR 20 – 61 years. We do not have epidemiologic information on the association of these individuals with students, so it is not possible to determine the circumstances of transmission. It is also possible that student status was misclassified for these individuals. This would result in an over-estimation of the amount of spread from outbreaks among students. It is also possible that some of these individuals were campus faculty or staff and therefore had differential exposure compared to the broader community.

**Figure 3:**
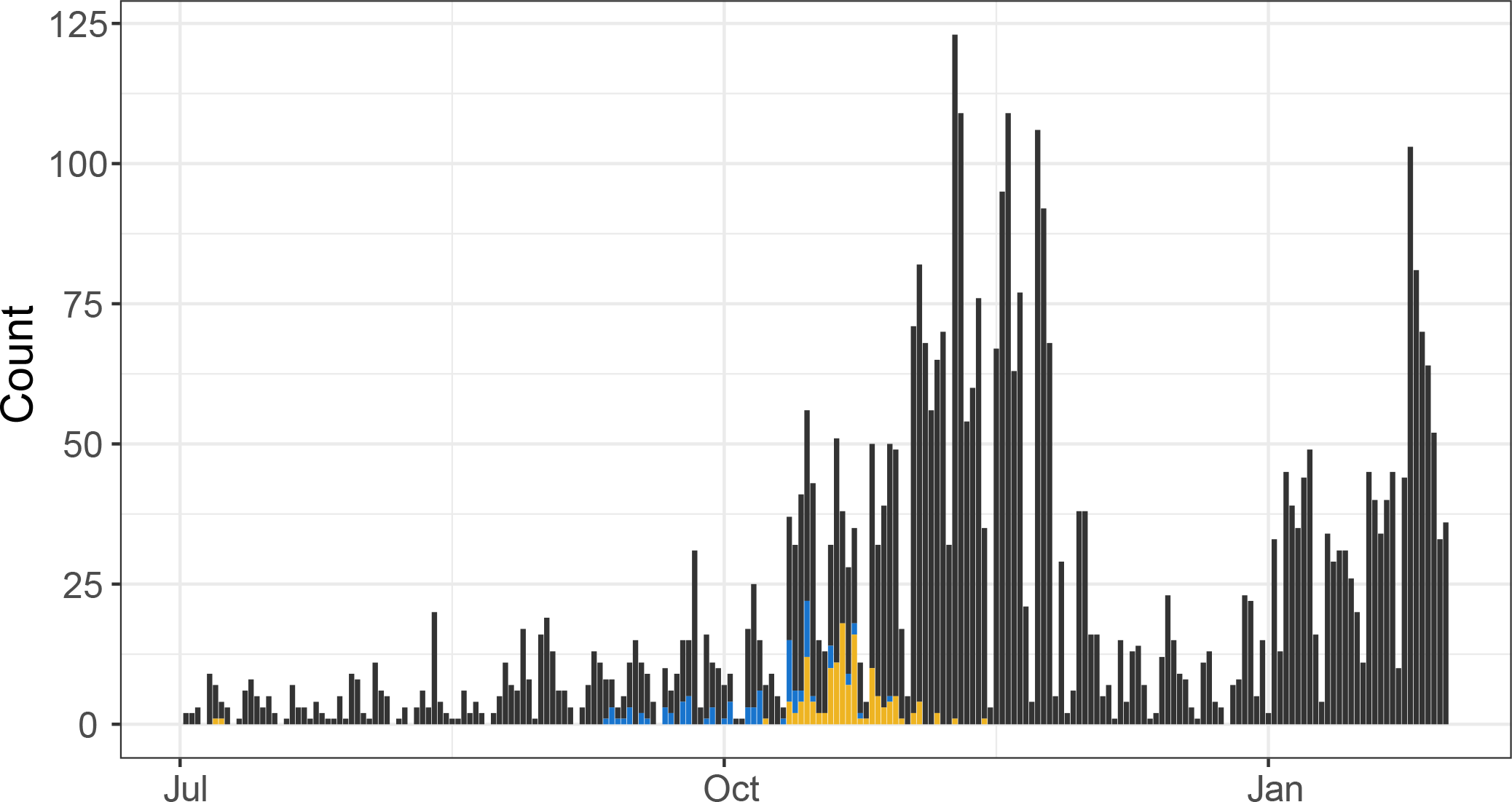
All genomes from Michigan available on GISAID from July 2020 – January 2021. Count of genomes is shown on the y-axis with time on the x-axis (binwidth = 1 day). Genomes from lineage B.1.593 are shown in blue, genomes from B.1.1.304 in yellow, and genomes from all other lineages are shown in grey.

**Figure 4:**
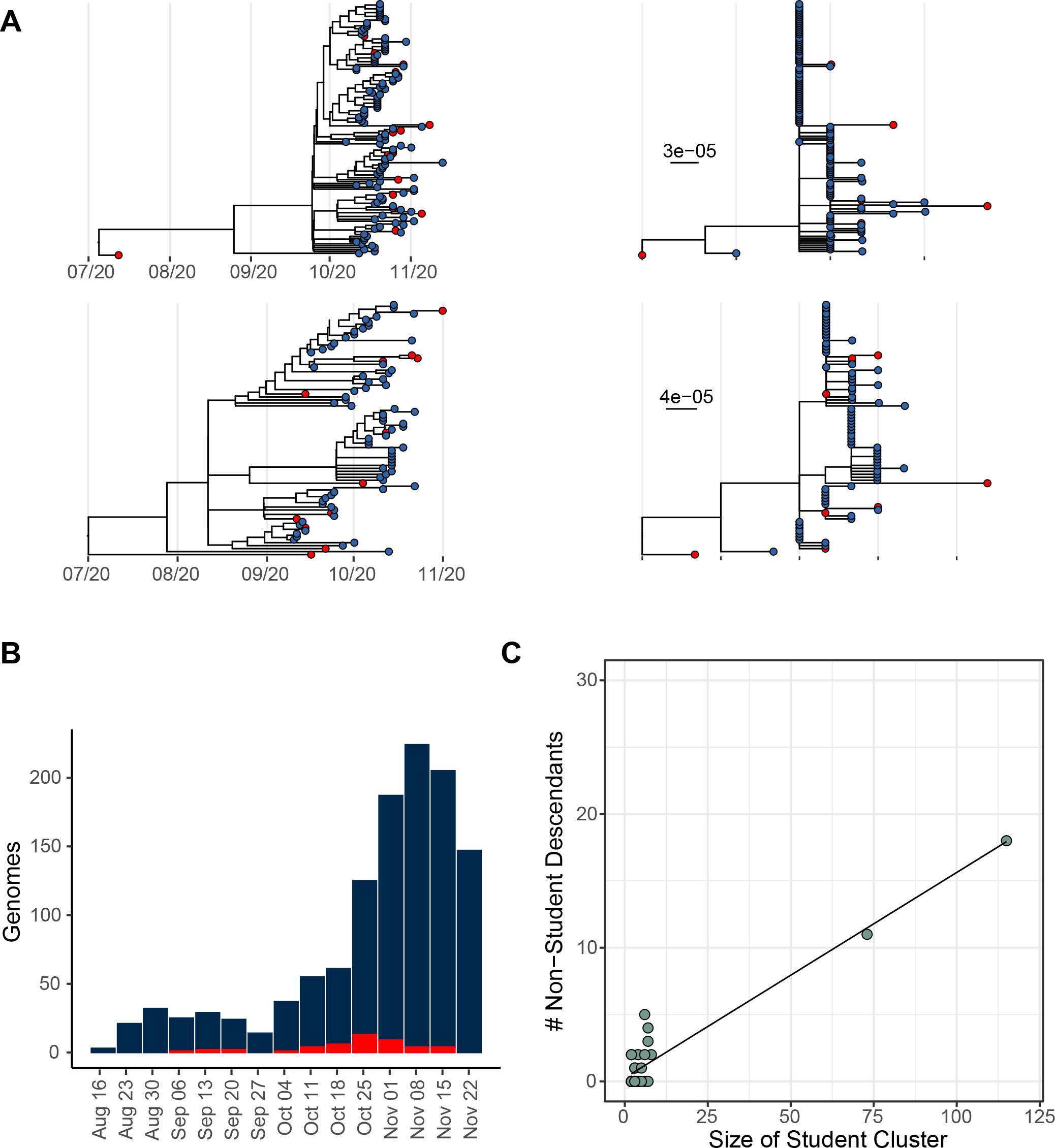
Spillover from student-associated clusters. (A) Maximum likelihood phylogenetic trees of student Cluster A (top) and Cluster B (bottom). Time-calibrated trees are displayed on the left and divergence trees on the right. Tip colors reflect genomes from students (light blue) and non-students (red). (B) Bar plot of the number of non-student genomes sequenced (y-axis, n = 1191) per week over the fall term (x-axis). Genomes that are derived from inferred “student” nodes are shown in red and genomes not derived from “student” nodes in dark blue. (C) For each non-singleton transmission lineage in students, the number of non-student descendants is shown (y-axis) by the number of students in the cluster (x-axis). A linear regression line is shown.

To further assess the spread from students to the community, we compared ancestral state models with different transition state matrices. Discrete trait reconstruction models that allowed for different transition rates between students and non-students exhibited better fit to the data compared to equal rates models (AIC 3228 vs. 3951, respectively), but a model that did not allow transitions from students to non-students exhibited a worse fit compared to the all-rates-different model (AIC 3805 vs. 3228, respectively). Overall, these results indicate that spillover from student-associated outbreaks into the broader community was infrequent and did not significantly contribute to the case increase in Washtenaw County in November and December 2020.

## Discussion

We conducted a prospective study of SARS-CoV-2 genomic surveillance focused on a large, public university and the surrounding community in southeastern Michigan. This is the first comprehensive characterization of SARS-CoV-2 genomic epidemiology across an entire university in the United States. A major strength of our study is high-density sampling of both the student population and the surrounding community by sequencing all available specimens from two major testing laboratories in Washtenaw County. This sampling strategy enabled us to evaluate the degree of crossover of SARS-CoV-2 transmission lineages between university students and the non-student population. These data illustrate the rapid transmission of COVID-19 within a large, public university population with remarkably little spread into the community.

We inferred over 200 unique introduction events of SARS-CoV-2 into the student population, demonstrating that the COVID-19 epidemic among students at the University of Michigan-Ann Arbor was not derived from a single introduction. Two of these introduced viruses resulted in rapid and sustained transmission for several weeks among students. As the semester progressed and community incidence rose, there was a larger influx of singleton introductions while the two large lineages declined. These results yield valuable insights into the spread of SARS-CoV-2 within a large university population. First, the majority of cases in students from September and October were derived from one of two dominant viral lineages, which co-existed for several weeks and circulated throughout several on-campus residences. This suggests uninterrupted spread among students that was not driven by associations with specific residence halls. We think it is unlikely that these two PANGO lineages (B.1.1.304 and B.1.593) have enhanced transmissibility or other notable intrinsic properties. These lineages have not disseminated widely as of this writing and do not exhibit the same mutations as other highly transmissible variants of concern. Second, these results show that a substantial fraction of cases in students (37%) were not related to other students (i.e., singleton introductions). This influx of newly introduced viruses late in the semester may have been driven by “outside-in” transmission from elsewhere in the community as incidence spiked in Washtenaw County. This means that in addition to preventing outbreaks among students at IHE, it is equally important to reduce overall county and regional COVID-19 incidence.

A key aspect of our study is the ability to estimate the extent to which outbreaks among students led to transmission into broader community. This question cannot be definitively answered through case count associations alone. We found very few cases in non-students that could be traced back genetically to clusters of students. Notably, the two PANGO lineages (B.1.1.304 and B.1.593) from the largest clusters among students have not persisted in Michigan since this time, providing additional confirmation that these outbreaks did not spur widespread transmission in the community. Of course, we did not sequence viruses from every infection and therefore cannot exclude that other spillover events may have occurred. Nevertheless, given our depth of sampling, it is unlikely that large outbreaks among students were the source of most COVID-19 infections in the community. If this was the case, we would expect to see vastly different patterns in the genomic data. In particular, we would expect to see a higher frequency of B.1.1.304 and B.1.593 in non-students from November and beyond. Although there are differences in context, setting, and mitigation measures here compared to other IHE, this study suggests that previous findings with limited community surveillance may generalize more broadly (Arnold et al., 2021; Currie et al., 2021; Weil et al., 2021).

There are other important limitations. First, we were not able to access specimens from commercial testing sites, and the number of detected transmission introductions is certainly an underestimate. It is likely that many singletons in our data are in fact linked to a small number of other student cases. It is also possible that there were differential contact patterns between students who obtained testing at Michigan Medicine sites and those who did not. Next, our study is not an epidemiologic investigation with contact tracing and individual behavioral information. It is difficult to reliably assess the effectiveness of any single mitigation measure that was implemented during the fall semester from these data alone. In particular, it is difficult to disentangle whether the lack of spread from students into the community is the result of university and county case identification and isolation measures for students, partitioning of social structures between IHE and the surrounding communities, or a combination of both. Thorough contact tracing investigations of IHE-associated outbreaks with dense genomic surveillance across populations may be able to resolve these questions in greater detail. It is also possible that these dynamics could have played out differently with earlier emergence of a highly transmissible variant, such as B.1.1.7 (Alpert et al., 2021; Washington et al., 2021). This work will be a valuable point of comparison for future studies examining the effects of more transmissible variants and vaccination on COVID-19 spread within IHE.

Our phylogenetic analysis of well-sampled genomic surveillance data provides insight into the spread of SARS-CoV-2 at a large, public university in the United States. There were hundreds of independent introductions into the student population, most of which did not result in detectable transmission in the local community. The small number of dominant lineages that circulated in the early and middle portions of the semester did not significantly contribute to the rise in county-level cases in November 2020. Nevertheless, we emphasize that even rare transmission events can have significant negative outcomes and disproportionally impact vulnerable populations, as evidenced by a recent study of an outbreak in a skilled nursing facility (Richmond et al., 2020). Additionally, SARS-CoV-2 infection can have severe clinical manifestations even in populations with generally lower risk (Logue et al., 2021; Tenforde, 2020). Therefore, it is critical that every effort is made by IHE and public health organizations to prevent and mitigate IHE-associated outbreaks in conjunction with measures at broader geographic levels.

## Methods

### Research Ethics and Sample Sources

We obtained residual upper respiratory diagnostic specimens that were positive for SARS-CoV-2 from the University of Michigan Clinical Microbiology Laboratory and the University Health Service. Use of residual SARS-CoV-2 positive specimens and collection of student status and on-campus residence were approved by the Institutional Review Board at the University of Michigan (Protocol HUM185966).

### Genome Amplification and Sequencing

We extracted RNA from nasopharyngeal specimens with the Invitrogen PureLink Pro 96 Viral RNA Mini kit, using 140 μL of input sample and eluting into 50 μL. We reverse transcribed RNA with SuperScript IV (ThermoFisher) by adding 1 μL of random hexamers and 1 μL of 10 mM dNTP to 11 μL of RNA extract, heating at 65 °C for 5 min, and placing on ice for 1 min. Then we added a reverse transcription master mix (4 μL of SuperScript IV buffer, 1 μL of 0.1M DTT, 1 μL of RNaseOUT RNase inhibitor, and 1 μL of SSIV enzyme) and incubated at 42 °C for 50 min, 70 °C for 10 min, and held at 4 °C. We amplified SARS-CoV-2 cDNA in two pools using the ARTIC Network version 3 primers. We used Q5 Hot Start High-Fidelity DNA Polymerase (NEB) and the following thermocycler protocol: 98 °C for 30 s, then 35 cycles of 98 °C for 15 s, 63 °C for 5 min, and final hold at 4 °C. We combined PCR products of the two primer pools in equal volumes for a given sample and purified the DNA product with 1X volume of AMPure beads (Beckman-Coulter). We prepared libraries for Illumina sequencing with the NEBNext Ultra II DNA Library Prep Kit (NEB) according to the manufacturer’s instructions. We pooled barcoded libraries for a batch of 96 samples by equal volume and gel extracted the band at 400-500 bp to remove adapter dimers. We quantified pooled libraries with the Qubit 1X dsDNA HS Assay Kit (ThermoFisher) and sequenced on an Illumina MiSeq (v2 chemistry, 2x250 cycles) at the University of Michigan Microbiome Core facility.

### Analysis of Raw Sequence Reads

We mapped reads to the Wuhan/Hu-1/2019 reference genome (GenBank MN908947.3) with BWA-MEM (Li, 2013). We trimmed the ARTIC amplification primer sequences with iVar 1.2.1 (Grubaugh et al., 2019). We determined consensus sequences with samtools mpileup and iVar 1.2.1. We assigned the consensus as the base with >50% frequency and placed an N at positions covered by fewer than 10 reads. Genomes with 29000 or more unambiguous bases (> 97%) were considered complete and used in downstream analysis. The consensus genomes are available on GISAID as listed in Supplemental Table 1.

### Case Definitions and Metadata

Of the samples from August 16 – November 24^th^ that yielded complete genomes, we used unique identifiers associated with each sample to obtain a single genome per individual. We used these identifiers to determine which individuals were students at the University of Michigan-Ann Arbor in the fall semester (undergraduate, graduate, or professional). We queried two databases hosted by the University Health Service to identify students and housing status. We identified 468 individuals who were listed as students in either or both databases. There were three individuals with conflicting classification; we investigated these further to determine the most accurate classification. Students with a campus residence hall listed in either database were considered “on-campus,” and the rest were considered “unknown.” We obtained case metrics in students from the University of Michigan COVID-19 Dashboard (https://campusblueprint.umich.edu/dashboard/, accessed on March 4^th^, 2021). We obtained data on new confirmed cases in Washtenaw County and Region 2S from the Michigan Department of Health and Human Services website (accessed March 4^th^, 2021).

### Phylogenetic Analysis and Discrete Trait Reconstruction

To provide phylogenetic context, we obtained a subsample of genomes on GISAID using the augur pipeline (Hadfield et al., 2018). We excluded genomes collected after 2020 and genomes with lengths less than 27000 bases. We aligned genomes with MAFFT as implemented in augur. We performed subsampling with priority on genomes with genetic similarity to Michigan sequences using the following rules: 100 genomes per month in Michigan, 5 genomes per month per division in the USA outside of Michigan, 1 genome per country per month in North America outside of the USA, and 2 genomes per country per month outside of North America. If not captured in the subsampling scheme, we added back the genomes presented here (n = 1659) as well as all other genomes from Michigan collected in August – December 2020. This resulted in an alignment with 7174 sequences, including 3318 genomes from Michigan, 2157 genomes from the USA outside of Michigan, and 1699 global genomes.

We masked the 5’ and 3’ ends of the alignment along with other sites commonly affected by sequencing errors and homoplasies. We inferred a maximum likelihood phylogeny with IQ-TREE with a GTR model and 1000 ultrafast bootstraps (Nguyen et al., 2015). We then used TreeTime to generate a time-scaled phylogeny rooted on Wuhan/Hu-1/2019 with a clock rate of 0.0008 with a standard deviation of 0.0004 substitutions/site/day (Sagulenko et al., 2018). We flagged and removed genomes that exceeded 3 interquartile ranges from the root-to-tip regression (n = 25, including two genomes we generated). We generated a new time-scaled phylogenetic tree with IQ-TREE and TreeTime as above with this filtered alignment (n = 7149). We used TempEst to fit and plot a root-to-tip regression of divergence over time (Rambaut et al., 2016). We used this tree as the basis for discrete-trait ancestral state reconstruction (ASR) using a binary model of student vs. non-student. We performed maximum likelihood ASR with TreeTime with a sampling bias of 2.5. We also performed Bayesian ASR using BEAST 1.10.4 using a fixed tree topology derived from TreeTime (Alpert et al., 2021; Drummond and Rambaut, 2007). We summarized the ASR results in R version 4.0.2. We inferred introductions from non-student to student populations at nodes with a BEAST probability of > 0.9. From these inferred “student” nodes, we counted the number of descendant student and non-student tips. Introduction nodes leading to only one student case were considered singletons, and introductions leading to two or more student cases were termed transmission lineages. To determine whether the contextual genomes were biasing our results, we generated a total of ten random subsamples of the global data using the schema described above and analyzed the data in the same manner (Figure S2C). Contextual genomes from GISAID used in these analyses are listed in Supplemental Table 1. We compared ancestral state models with different transition matrices using the functions implemented in the “castor” R package (Louca and Doebeli, 2018).

### Data and Code Availability

The consensus genomes we generated for this study are publicly available on GISAID. Accessions for these sequences as well as other genomes from GISAID used in these analyses are listed in Supplementary Table 1. Analysis code for the generation of consensus sequences and phylogenetic analysis are available on GitHub at the following link: https://github.com/lauringlab/SARS2_Fall_2020.

## Data Availability

The consensus genomes we generated for this study are publicly available on GISAID. Accessions for these sequences as well as other genomes from GISAID used in these analyses are listed in Supplementary Table 1. Analysis code for the generation of consensus sequences
and phylogenetic analysis are available on GitHub at the following link: https://github.com/lauringlab/SARS2_Fall_2020.

## Acknowledgements

We thank the University of Michigan Clinical Microbiology Laboratory, COVID-19 Central Biorepository, and University Health Service for sharing samples. We thank the University of Michigan Microbiome Core facility for their assistance in genome sequencing. We thank Teodora Jevtic for assistance in obtaining individual metadata. We thank Nathan Grubaugh, Anderson Brito, and Simon Dellicour for helpful discussion on phylogenetic analysis. This work was funded by CDC contract 75D30120C09870.

## Supplemental Figure Legends

**Supplemental Figure 1:**
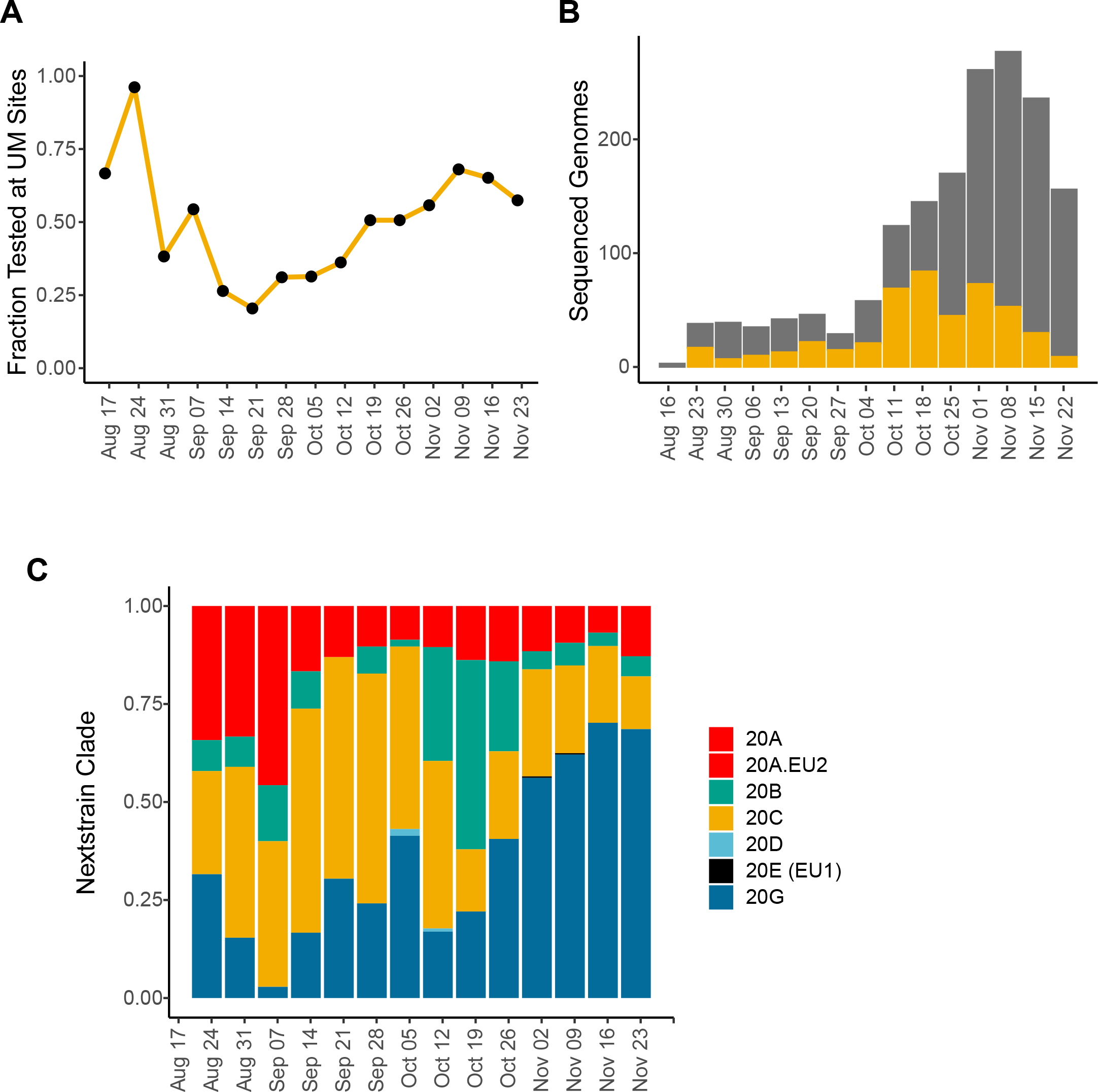
Information on sampling density and sequenced genomes. (A) Fraction of cases in students that were tested at University of Michigan testing sites (y-axis) per week over the fall term (x-axis). (B) Number of complete genomes sequenced (y-axis) per week of the fall term (x-axis). The number of genomes from University of Michigan students is shown in yellow. (C) Stacked bar plot of genomes sequenced (y-axis) per week over the fall term (x-axis). Bar colors represent the Nextstrain clades of each genome.

**Supplemental Figure 2:**
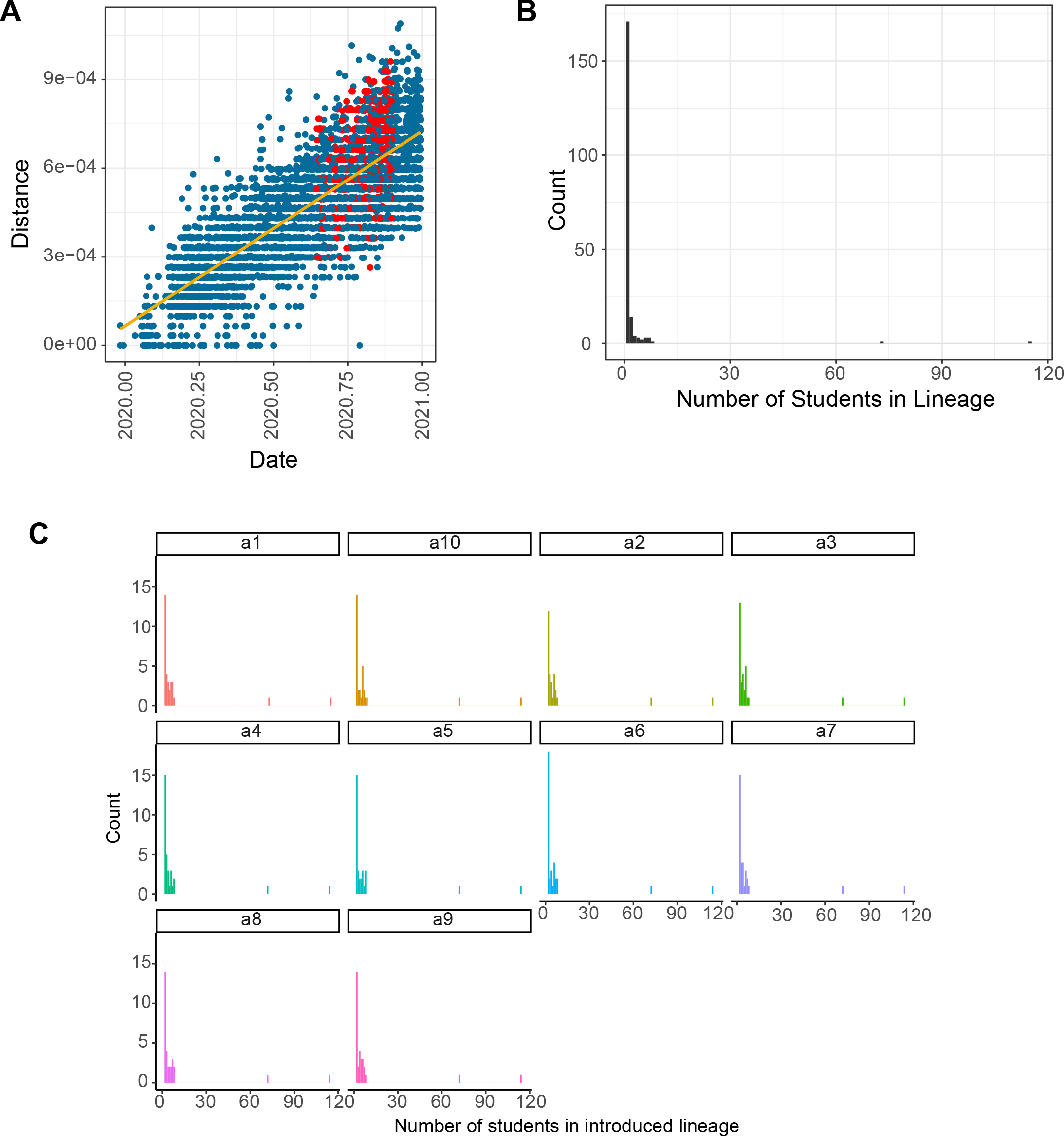
Information on phylogenetic analysis of introductions into student population. (A) Root-to-tip divergence of all 7149 genomes in the analysis presented in Figures 2 and 3, with genetic distance (y-axis) displayed by time (x-axis). Genomes sequenced are shown in red and others are shown in blue. A linear regression line is shown in yellow. (B) Histogram of the number of students in each transmission lineage (bin width = 1). (C) Histogram of the number of students in each non-singleton transmission lineage as in (B), with each facet showing analysis for ten independent subsamples of contextual genomes as described in the text.

